# Epidemiology of post-COVID syndrome following hospitalisation with coronavirus: a retrospective cohort study

**DOI:** 10.1101/2021.01.15.21249885

**Authors:** Daniel Ayoubkhani, Kamlesh Khunti, Vahé Nafilyan, Thomas Maddox, Ben Humberstone, Sir Ian Diamond, Amitava Banerjee

## Abstract

**Objectives:** The epidemiology of post-COVID syndrome (PCS) is currently undefined. We quantified rates of organ-specific impairment following recovery from COVID-19 hospitalisation compared with those in a matched control group, and how the rate ratio (RR) varies by age, sex, and ethnicity.

**Design:** Observational, retrospective, matched cohort study.

**Setting:** NHS hospitals in England.

**Participants:** 47,780 individuals (mean age 65 years, 55% male) in hospital with COVID-19 and discharged alive by 31 August 2020, matched to controls on demographic and clinical characteristics.

**Outcome measures:** Rates of hospital readmission, all-cause mortality, and diagnoses of respiratory, cardiovascular, metabolic, kidney and liver diseases until 30 September 2020.

**Results:** Mean follow-up time was 140 days for COVID-19 cases and 153 days for controls. 766 (95% confidence interval: 753 to 779) readmissions and 320 (312 to 328) deaths per 1,000 person-years were observed in COVID-19 cases, 3.5 (3.4 to 3.6) and 7.7 (7.2 to 8.3) times greater, respectively, than in controls. Rates of respiratory, diabetes and cardiovascular events were also significantly elevated in COVID-19 cases, at 770 (758 to 783), 127 (122 to 132) and 126 (121 to 131) events per 1,000 person-years, respectively. RRs were greater for individuals aged <70 than ≥ 70 years, and in ethnic minority groups than the White population, with the biggest differences observed for respiratory disease: 10.5 [9.7 to 11.4] for <70 years versus 4.6 [4.3 to 4.8] for ≥ 70 years, and 11.4 (9.8 to 13.3) for Non-White versus 5.2 (5.0 to 5.5) for White.

**Conclusions:** Individuals discharged from hospital following COVID-19 face elevated rates of multi-organ dysfunction compared with background levels, and the increase in risk is neither confined to the elderly nor uniform across ethnicities. The diagnosis, treatment and prevention of PCS require integrated rather than organ- or disease-specific approaches. Urgent research is required to establish risk factors for PCS.

## Introduction

In the early coronavirus disease (COVID-19) pandemic in the UK, approximately 6% of people were thought to have had severe acute respiratory syndrome coronavirus 2 (SARS-CoV2) infection, rising to 13% in London.[1] The majority of the research, health service and media response to the virus has focused on direct (through infection) and indirect (through impact on individual behaviours and health system changes) effects of COVID-19 on mortality[2] and short-term impacts.[3,4] However, the longer-term effects on morbidity need to be studied in order to plan for effective healthcare delivery and capacity.

Since SARS-CoV2 infection was recognised in late 2019, academic and clinical emphasis has been on respiratory manifestations.[5] However, there is increasing evidence for direct multi-organ effects,[6-10] as well as indirect effects on other organ systems and disease processes, such as cardiovascular diseases and cancers, through changes in healthcare delivery and patient behaviours.[11-13] Although the long-term impact of COVID-19 on individuals and health systems is becoming clear, there is an urgent need for investigation across organ systems.

Long COVID, or post-COVID syndrome (PCS), is not a single condition, and has been defined by the National Institute for Health and Care Excellence (NICE) as “signs and symptoms that develop during or following an infection consistent with COVID-19 which continue for more than 12 weeks and are not explained by an alternative diagnosis.”[14] Guidelines recommend referral to PCS assessment clinics if post-COVID symptoms persist for 6-12 weeks.[14] Pre-existing conditions and risk factors are predictors of acute COVID-19 outcomes (such as intensive care admissions and mortality[2]), but the epidemiology of PCS has been less well defined[15,16] due to unclear medium- and long-term pathophysiology across organ systems. As PCS clinics are implemented, characterisation of disease epidemiology will aid appropriate diagnosis, care, public health interventions and policy, and resource planning.

The existing evidence base shows large variations in estimates of PCS prevalence and incidence due to differences in study populations, recruitment methods, follow-up periods, and sample sizes. Most studies to date have concentrated on symptoms associated with PCS rather than organ impairment, and few have made use of a control group permitting the inference of counterfactual outcomes. Therefore, using national electronic health records and death registrations for individuals in England, we quantified the incidence of mortality, health service utilisation, and organ-specific impairment following discharge from hospital with COVID-19. We estimated rate ratios of post-discharge adverse events compared with those in a matched control group, and heterogeneity in this rate ratio across demographic groups.

## Methods

### Study design and data sources

This was an observational, retrospective, matched cohort study of individuals in hospital with COVID-19 using Hospital Episode Statistics (HES) Admitted Patient Care (APC) records for England until 31 August 2020. The study also utilised the General Practice Extraction Service (GPES) Data for Pandemic Planning and Research (GDPPR) dataset, an extract of approximately 35,000 clinical codes for over 56 million individuals registered at General Practices in England, until 30 September 2020. Death registrations data from the Office for National Statistics (ONS) were linked for deaths until 30 September 2020 and registered by 7 October 2020.

### Study population

Individuals with COVID-19 were included in the study if they had a hospital episode starting from 1 January 2020 and ending by 31 August 2020 with a primary diagnosis of COVID-19, identified using International Statistical Classification of Diseases and Related Health Problems 10th Revision (ICD-10) codes U07.1 (virus identified) and U07.2 (virus not identified); that is, by a positive laboratory test or clinical diagnosis. Individuals with COVID-19 were excluded if they had not been discharged alive by 31 August 2020 or had an unknown date of birth and/or sex. The index date was set to date of discharge following the first hospital episode with COVID-19 as the primary diagnosis.

Candidate controls comprised individuals who: (i) did not meet the COVID-19 inclusion criteria specified above; (ii) had at least one GDPPR record between 1 January 2019 (one year before the start of the follow-up period) and 30 September 2020 (the end-of-study date); and (iii) had not died before 1 January 2020. Each control had the same index date as their matched COVID-19 case.

### Outcome variables

All individuals were followed-up from index date to 30 September 2020 or date of death (whichever was earlier) for the following adverse events: all-cause mortality; hospital readmission for any reason; respiratory disease; major adverse cardiovascular event (MACE, a composite of heart failure [HF], myocardial infarction [MI], stroke, and arrhythmia); diabetes (type 1 or 2); chronic kidney disease (CKD) stages 3-5 (including dialysis and kidney transplant); and chronic liver disease (CLD).

Respiratory, MACE, diabetes, CKD and CLD events were identified from diagnoses made in primary care and in hospital (using only primary ICD-10 codes for the latter), except for the arrythmia component of MACE for which primary care data were not available.

### Matching variables

COVID-19 cases were matched to controls on baseline demographic and clinical characteristics that may confound the relationship between COVID-19 and outcomes (Supplementary Table 1), established over the ten-year look-back period 1 January 2010 to 31 December 2019. Comorbidities were identified from diagnoses made in primary care and in hospital (using both primary and secondary ICD-10 codes for the latter).

### Statistical techniques

Distributions of baseline characteristics were compared between individuals with COVID-19 and a random 0.5% sample of the general population using Chi squared tests and standardised differences in proportions, where a standardised difference in excess of 10% was taken as evidence of large imbalance between the groups.[17]

COVID-19 cases and controls were matched using exact 1:1 matching. Matched pairs were discarded if the control died before the corresponding COVID-19 case’s index date. All covariates were discretised prior to matching, including an ‘Unknown’ category comprising individuals with missing values.

We computed rates of adverse events per 1,000 person-years of exposure time in COVID-19 cases and controls, and derived rate ratios contrasting these event rates, to estimate the ‘average treatment effect on the treated.’ 95% confidence intervals (CIs) were estimated using the Poisson distribution.

We estimated events rates for the full study population firstly by using all adverse events, and secondly by considering only new-onset cases (that is, where the individual did not have a diagnosis for the condition over the ten-year look-back period). All-diagnoses event rates were stratified by sex, coarse age group (<70 years, ≥ 70 years), and coarse ethnic group (White, Non-White). In secondary analysis, individuals with COVID-19 were stratified according to whether they were admitted to an intensive care unit (ICU) during their hospital spell as a measure of disease severity. All statistical analyses were conducted using R version 4.0.2.

## Results

### Study participants

Of 86,955 individuals in hospital with COVID-19 during the study period, 53,795 (61.9%) had been discharged alive by end-of-study (Figure 1). After excluding individuals with unknown age or sex and those who could not be matched to a control, 47,780 COVID-19 cases (4,745 ICU and 43,035 non-ICU) were included in the analysis, representing 90.8% of those discharged alive with known age and sex. Mean follow-up times were 140 days (standard deviation [SD] 50 days, maximum 253 days) for COVID-19 cases and 153 days (SD 33 days, maximum 253 days) for controls.

**Figure 1.**
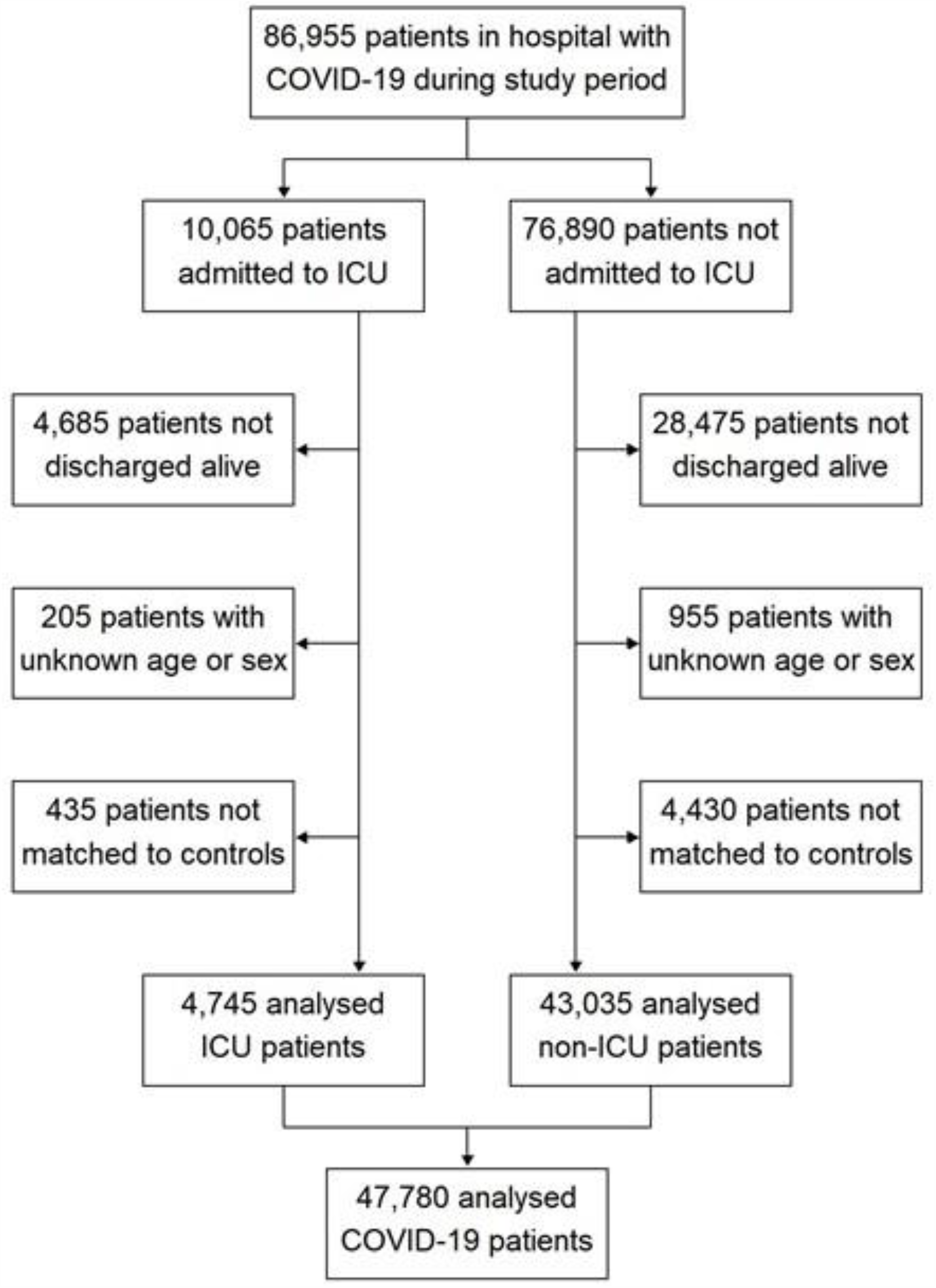
Study population flow diagram.

At baseline, individuals with COVID-19 had a mean age of 64.5 years (SD 19.2 years) and 54.9% were male. Compared with the general population, individuals in hospital with COVD-19 were more likely to be aged ≥ 50 years, male, living in a deprived area, a former smoker, and overweight or obese (Table 1). Individuals with COVID-19 were also more likely to be comorbid than the general population, with a higher prevalence of prior hospitalisation and all measured pre-existing conditions (most notably hypertension, MACE, respiratory disease and diabetes).

**Table 1.** Baseline characteristics of individuals in hospital with COVID-19 in England compared with those of a random sample from the general population

| Characteristic | Category | COVID-19 cases<br>(n = 47,780) | General population<br>(n = 239,380) | P-value | Standardised<br>difference (%) |
| --- | --- | --- | --- | --- | --- |
| Age | <30 years | 2,255 (4.7%) | 75,700 (31.6%) | <0.0001 | -74.5 |
|  | 30-49 years | 7,760 (16.2%) | 61,430 (25.7%) | <0.0001 | -23.3 |
|  | 50-69 years | 15,945 (33.4%) | 61,500 (25.7%) | <0.0001 | 16.9 |
|  | ≥70 years | 21,825 (45.7%) | 35,715 (14.9%) | <0.0001 | 71.0 |
| Sex | Male | 26,245 (54.9%) | 107,890 (45.1%) | <0.0001 | 19.8 |
|  | Female | 21,535 (45.1%) | 126,450 (52.8%) | <0.0001 | -15.5 |
| Ethnicity | White | 34,355 (71.9%) | 151,180 (63.2%) | <0.0001 | 18.8 |
|  | Asian | 4,320 (9.0%) | 15,150 (6.3%) | <0.0001 | 10.2 |
|  | Black | 2,565 (5.4%) | 6,840 (2.9%) | <0.0001 | 12.7 |
|  | Mixed/Other | 1,430 (3.0%) | 7,010 (2.9%) | 0.4731 | 0.4 |
|  | Unknown | 5,110 (10.7%) | 59,205 (24.7%) | <0.0001 | -37.4 |
| IMD quintile | 1 (most deprived) | 11,510 (24.1%) | 48,555 (20.3%) | <0.0001 | 9.2 |
|  | 2 | 10,970 (23.0%) | 47,680 (19.9%) | <0.0001 | 7.4 |
|  | 3 | 9,265 (19.4%) | 47,125 (19.7%) | 0.1340 | -0.8 |
|  | 4 | 8,315 (17.4%) | 46,040 (19.2%) | <0.0001 | -4.7 |
|  | 5 (least deprived) | 7,695 (16.1%) | 44,795 (18.7%) | <0.0001 | -6.9 |
|  | Unknown | 25 (<0.1%) | 5,185 (2.2%) | <0.0001 | -20.3 |
| Smoking status | Current | 4,000 (8.4%) | 38,040 (15.9%) | <0.0001 | -23.2 |
|  | Former | 19,560 (40.9%) | 56,210 (23.5%) | <0.0001 | 38.0 |
|  | Never | 20,295 (42.5%) | 93,750 (39.2%) | <0.0001 | 6.7 |
|  | Unknown | 3,920 (8.2%) | 51,375 (21.5%) | <0.0001 | -38.0 |
| BMI | <25 kg/m<sup>2</sup> | 9,415 (19.7%) | 60,140 (25.1%) | <0.0001 | -13.0 |
|  | 25 to <30 kg/m<sup>2</sup> | 12,140 (25.4%) | 48,290 (20.2%) | <0.0001 | 12.5 |
|  | ≥30 kg/m<sup>2</sup> | 15,390 (32.2%) | 40,795 (17.0%) | <0.0001 | 35.8 |
|  | Unknown | 10,835 (22.7%) | 90,155 (37.7%) | <0.0001 | -33.1 |
| Previous hospitalisation |  | 39,575 (82.8%) | 150,510 (62.9%) | <0.0001 | 46.0 |
| Hypertension |  | 24,720 (51.7%) | 43,170 (18.0%) | <0.0001 | 75.6 |
| Respiratory disease |  | 19,440 (40.7%) | 38,695 (16.2%) | <0.0001 | 56.5 |
| Asthma |  | 8,695 (18.2%) | 27,345 (11.4%) | <0.0001 | 19.2 |
| COPD |  | 6,565 (13.7%) | 7,090 (3.0%) | <0.0001 | 39.7 |
| Other |  | 11,890 (24.9%) | 20,405 (8.5%) | <0.0001 | 45.0 |
| Diabetes |  | 11,680 (24.4%) | 16,670 (7.0%) | <0.0001 | 49.5 |
| Type 1 |  | 1,235 (2.6%) | 1,770 (0.7%) | <0.0001 | 14.5 |
| Type 2 |  | 11,530 (24.1%) | 15,810 (6.6%) | <0.0001 | 50.1 |
| MACE |  | 11,650 (24.4%) | 13,385 (5.6%) | <0.0001 | 54.6 |
| Heart failure |  | 5,255 (11.0%) | 4,150 (1.7%) | <0.0001 | 38.7 |
| Stroke |  | 3,040 (6.4%) | 3,100 (1.3%) | <0.0001 | 26.6 |
| Myocardial infarction |  | 2,265 (4.7%) | 3,160 (1.3%) | <0.0001 | 20.1 |
| Arrhythmia |  | 7,170 (15.0%) | 7,540 (3.1%) | <0.0001 | 42.2 |
| Cancer |  | 9,820 (20.5%) | 22,090 (9.2%) | <0.0001 | 32.2 |
| CKD stages 3-5 |  | 6,075 (12.7%) | 7,930 (3.3%) | <0.0001 | 35.1 |
| Dementia |  | 5,010 (10.5%) | 2,560 (1.1%) | <0.0001 | 41.2 |
| Osteoporosis |  | 4,390 (9.2%) | 5,175 (2.2%) | <0.0001 | 30.7 |
| Rheumatoid arthritis |  | 1,890 (4.0%) | 2,650 (1.1%) | <0.0001 | 18.2 |
| Chronic liver disease |  | 1,380 (2.9%) | 3,005 (1.3%) | <0.0001 | 11.5 |

Table notes: BMI: body mass index; CKD: chronic kidney disease; COPD: chronic obstructive pulmonary disease; IMD: Index of Multiple Deprivation; MACE: major adverse cardiovascular event. Baseline characteristics extracted from primary care records and hospital episodes over the period 1 January 2010 to 31 December 2019. The general population is a random 0.5% sample of individuals with primary care records between 1 January 2019 and 30 September 2020.

### Rates of adverse events in individuals with COVID-19 following discharge

Of 47,780 individuals in hospital with COVID-19 over the study period, 29.4% were re-admitted and 12.3% died following discharge (Table 2). These events occurred at rates of 766 (CI: 753 to 779) readmissions and 320 (312 to 328) deaths per 1,000 person-years, which were 3.5 (3.4 to 3.6) and 7.7 (7.2 to 8.3) times greater, respectively, than those in matched controls. Respiratory disease was diagnosed in 14,140 individuals (29.6%) following discharge, with 6,085 of these being new-onset cases; the resulting event rates of 770 (758 to 783) and 539 (525 to 553) per 1,000 person-years, respectively, were 6.0 (5.7 to 6.2) and 27.3 (24.0 to 31.2) times greater than those in controls.

**Table 2.** Counts and rates of death, readmission and respiratory disease contrasting individuals with COVID-19 in England discharged from hospital by 31 August 2020 with matched controls

| Adverse event<br>(sample size per group) | COVID-19 cases |  | Control group |  |
| --- | --- | --- | --- | --- |
|  | Events<br>(n, %) | Rate per 1,000 person-years (95% CI) | Events<br>(n, %) | Rate per 1,000 person-years (95% CI) |
| Death<br>(n = 47,780) | 5,875<br>(12.3%) | 320.0<br>(311.9 to 328.3) | 830<br>(1.7%) | 41.3<br>(38.6 to 44.3) |
| Readmission to hospital<br>(n = 47,780) | 14,060<br>(29.4%) | 766.0<br>(753.4 to 778.8) | 4,385<br>(9.2%) | 218.9<br>(212.4 to 225.4) |
| Respiratory disease (all events)<br>(n = 47,780) | 14,140<br>(29.6%) | 770.5<br>(757.8 to 783.3) | 2,585<br>(5.4%) | 129.1<br>(124.2 to 134.2) |
| Respiratory disease (new onset)<br>(n = 28,335) | 6,085<br>(21.5%) | 538.9<br>(525.5 to 552.6) | 240<br>(0.8%) | 19.7<br>(17.3 to 22.4) |

Post-discharge diagnoses of diabetes, MACE, CKD and CLD were made for 4.9%, 4.8%, 1.5% and 0.3% of individuals with COVID-19, respectively, occurring at rates of 127 (122 to 132) diabetes, 126 (121 to 131) MACE, 39 (36 to 42) CKD and 7 (6 to 9) CLD events per 1,000 person-years (Figure 2). A similar pattern was observed when only new-onset cases were considered, but at lower rates of 29 (26 to 32) diabetes, 66 (62 to 70) MACE, 15 (13 to 17) CKD and 4 (3 to 5) CLD events per 1,000 person-years. Those with COVID-19 received post-discharge diagnoses of MACE, CLD, CKD and diabetes 3.0 (2.7 to 3.2), 2.8 (2.0 to 4.0), 1.9 (1.7 to 2.1) and 1.5 (1.4 to 1.6) times more frequently, respectively, than in the matched control group. See Supplementary Table 2 for detailed results.

**Figure 2.**
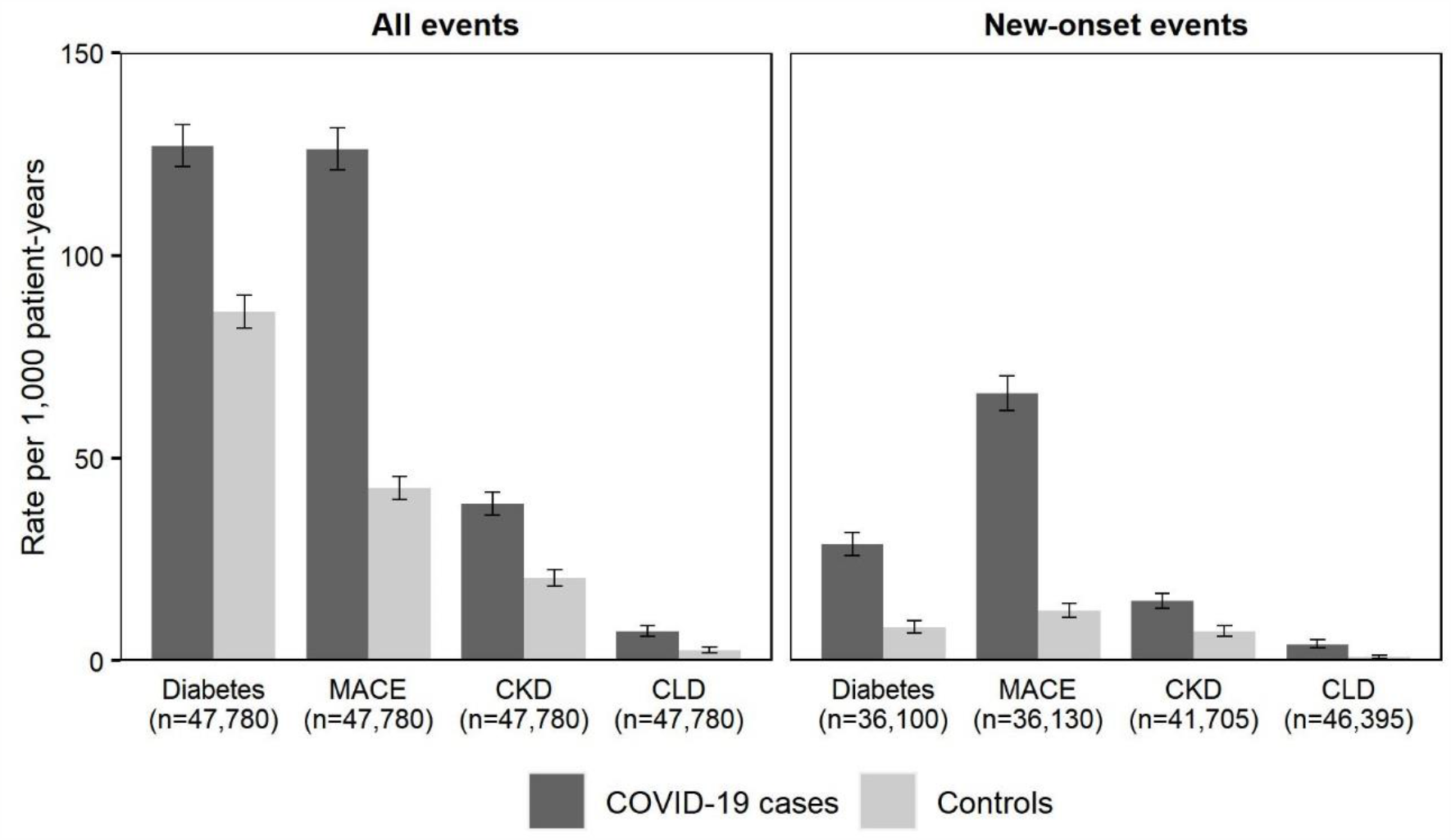
Rates of adverse events contrasting individuals with COVID-19 in England discharged from hospital by 31 August 2020 with matched controls Figure notes: CKD: chronic kidney disease stages 3-5; CLD: chronic liver disease; MACE: major adverse cardiovascular event. Adverse events calculated from hospital episodes to 31 August 2020, and primary care records and deaths registrations to 30 September 2020. COVID-19 cases were matched to controls on baseline demographic characteristics (age, sex, ethnicity, region, Index of Multiple Deprivation quintile, smoking status) and clinical histories (hypertension, MACE, respiratory disease, CKD, CLD, diabetes, cancer).

In secondary analysis, rates of post-discharge adverse events remained significantly elevated among individuals with COVID-19 compared with matched controls after stratifying by ICU versus non-ICU admission (Supplementary Table 3). Individuals requiring ICU admission experienced greater rates of post-discharge respiratory disease and diabetes than those not in ICU, but the opposite was true for rates of death, readmission and MACE.

### Rate ratios of adverse events across demographic characteristics

Rates of all post-discharge adverse events were greater in individuals with COVID-19 aged ≥ 70 years than <70 years, while rates of all events other than diabetes were greater in the White ethnic group than the Non-White group (Supplementary Table 4). However, the rate ratio of adverse events (contrasting COVID-19 cases and matched controls) was greater in individuals aged <70 years than ≥ 70 years for all event types (Figure 3), with the biggest differences in rate ratios being observed for death (14.1 [11.0 to 18.3] for <70 years versus 7.7 [7.1 to 8.3] for ≥ 70 years) and respiratory disease (10.5 [9.7 to 11.4] for <70 years versus 4.6 [4.3 to 4.8] for ≥ 70 years). Ethnic differences in rate ratios were most pronounced for respiratory disease, at 11.4 (9.8 to 13.3) for individuals in the Non-White group compared with 5.2 (5.0 to 5.5) in the White group. Differences in rate ratios of adverse events between males and females were generally small. See Supplementary Table 4 for detailed results.

**Figure 3.**
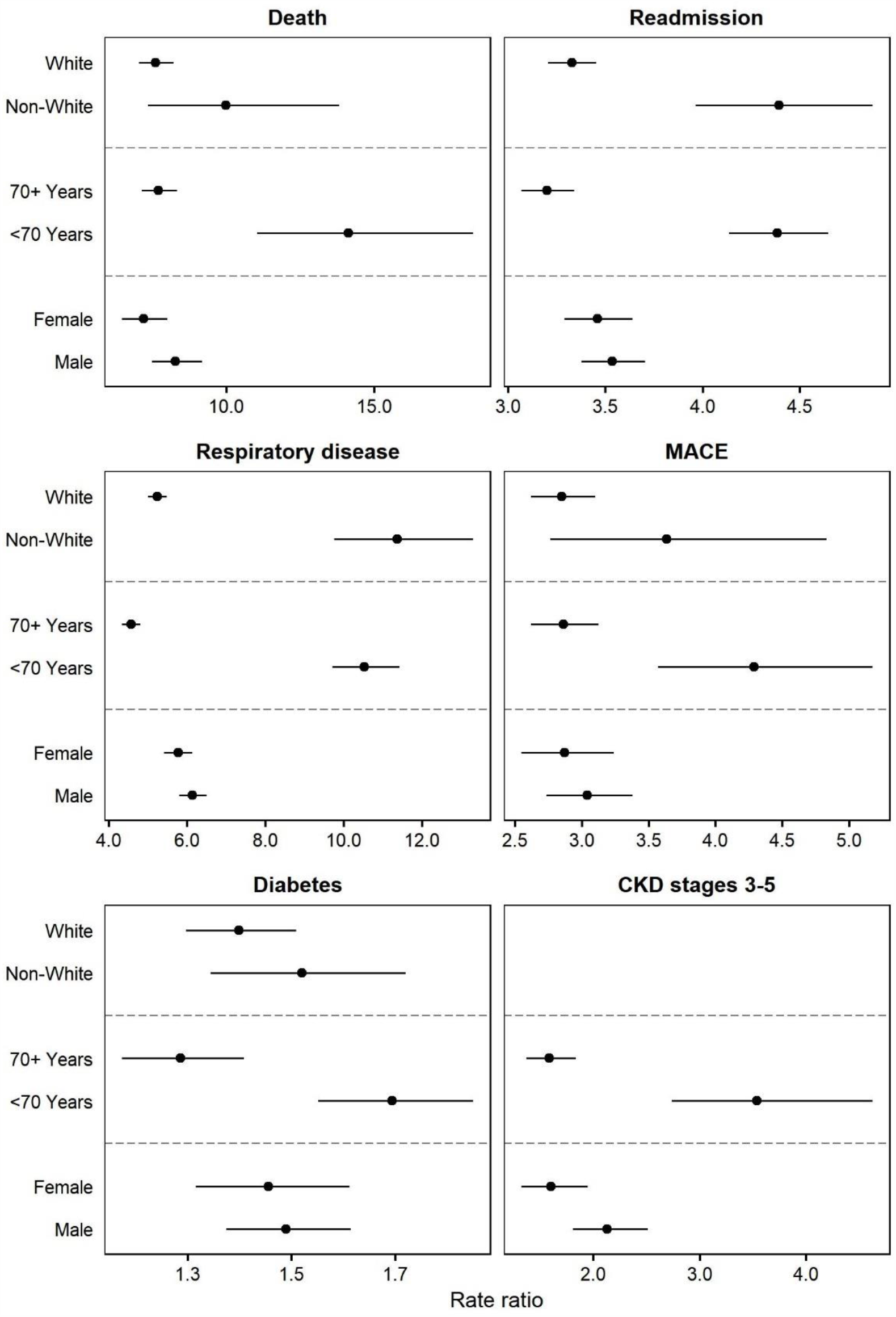
Rate ratios of adverse events contrasting individuals with COVID-19 in England discharged from hospital by 31 August 2020 with matched controls, stratified by demographic factors Figure notes: CKD: chronic kidney disease; MACE: major adverse cardiovascular event. Adverse events calculated from hospital episodes to 31 August 2020, and primary care records and deaths registrations to 30 September 2020. COVID-19 cases were matched to controls on baseline demographic characteristics (age, sex, ethnicity, region, Index of Multiple Deprivation quintile, smoking status) and clinical histories (hypertension, MACE, respiratory disease, CKD, CLD, diabetes, cancer). Rate ratios for CKD could not be stratified by ethnic group due to insufficient event counts in the control group.

## Discussion

### Principal findings

In the largest study to-date to examine PCS in individuals hospitalised with COVID-19, comprising 47,780 COVID-19 cases with matched controls, we describe three major findings. Firstly, COVID-19 hospitalisation was associated with increased risk of readmission and death following discharge, relative to that in individuals of similar demographic and clinical profiles over the same period; nearly a third of people post COVID-19 hospital discharge were re-admitted and more than 1 in 10 died.

Secondly, rates of post-discharge multi-organ dysfunction were elevated in individuals with COVID-19 compared with those in the matched control group, suggesting extrapulmonary pathophysiology. Diabetes and MACE were particularly common, both when considering all post-discharge events (which may reflect a combination of new-onset cases and exasperation of pre-existing conditions) and only incident cases.

Finally, the absolute risk of post-discharge adverse events was greater for individuals aged ≥ 70 years than <70 years, and for individuals of White ethnic background than in the Non-White group. However, when contrasted against the background rates of adverse events that might be expected to occur in these groups in the general population, younger and ethnic minority individuals faced greater relative risks than those aged ≥ 70 years and those in the White group, respectively.

### Comparison with related studies

Our results are consistent with hypothesised biological mechanisms associated with respiratory,[18] cardiovascular,[19] metabolic,[20] renal,[10] and hepatic[8] involvement in COVID-19, and extend the early evidence base surrounding PCS which has been described as being “limited” and generally of low quality.[21]

In a more recent study of 1,775 US veterans hospitalised with COVID-19, 20% were readmitted and 9% died within 60 days of discharge;[22] after restricting follow-up time in our study to the same duration, we found similar prevalence rates of 23% and 9%, respectively. The US study did not analyse organ-specific endpoints and was conducted in a specific population; we can thus now add that COVID-19 is associated with post-discharge manifestations in a range of organs in the general population.

Multi-organ involvement following COVID-19 infection was detected in 201 low-risk individuals in the UK (18% hospitalised with COVID-19), with impairment of the lungs (33%), heart (32%), kidneys (12%) and liver (10%) found to be common.[23] These prevalence rates are higher than those estimated in our study, though the degree of organ impairment was generally found to be mild and therefore likely to be subclinical.

In a case series of 213 discharged individuals with COVID-19 in the US, 10% were re-admitted and 2% died over a median follow-up time of 80 days,[24] compared with our much greater estimates of 29% and 12%, respectively (but over a longer median follow-up time of 160 days). However, the small sample size precludes extrapolation to broader populations.

COVID-19 was associated with increased odds of acute kidney injury, renal replacement therapy, insulin use, pulmonary embolism, stroke, myocarditis, arrythmia, and elevated troponin in US veterans hospitalised with COVID-19 versus a control of seasonal influenza.[25] The index event was admission rather than discharge, so the results do not necessarily capture long-term outcomes of COVID-19, but they are indicative of physiological changes in multiple organs at the acute phase of the disease and therefore support our own findings.

Pulmonary lesions were found in hospitalised COVID-19 patients in Wuhan, China, though only at a short follow-up duration of three weeks post-discharge.[26] Cardiovascular magnetic resonance (CMR) imaging revealed myocardial inflammation in German participants who recovered from acute COVID-19,[6] while myocarditis was also detected by CMR imaging in post-acute US college athletes.[27] These studies are suggestive of pulmonary and myocardial involvement in individuals with COVID-19 and, although small sample sizes and highly specific study populations make it difficult to generalise the results, they shed some light on possible pathophysiological mechanisms that underly our own findings.

### Implications of findings

With over 3 million people in the UK having tested positive for COVID-19 at the time of writing,[28] and many more who had the disease but never received a test, our findings suggest that the long-term burden of COVID-related morbidity on hospitals and broader healthcare systems is likely to be substantial. PCS comes on a backdrop of healthcare challenges, particularly sustainable high-quality care for long-term conditions: inequalities in health, access and provision; incomplete pathways across community and hospital care; inadequate research translation to clinical practice; and insufficient resources. Our findings across organ systems suggest that the diagnosis, treatment and prevention of PCS will require integrated rather than organ- or disease-specific approaches. Integrated care pathways are structured, multidisciplinary care plans for specific conditions,[29] which have been effective in other diseases such as chronic obstructive pulmonary disease and may have utility in the management of PCS.

### Strengths and limitations

The main strength of our study lies in its size and completeness, as it includes all individuals in England in hospital with COVID-19 observed over a follow-up period of up to several months. Use of a matched control group allowed rates of post-discharge adverse events in individuals with COVID-19 to be compared against counterfactual outcomes – what might have been observed given the background risk in these individuals.

Like all observational studies, we cannot rule out the possibility of residual confounding (for example, due to biomarkers or socio-economic exposures omitted from our matching set) which precludes our ability to draw definitive causal conclusions. The limited number of events in the control group meant we were unable to disaggregate rate ratios stratified by age and ethnicity beyond broad ‘<70 years verses ≥ 70 years’ and ‘White versus Non-White’ comparisons, despite the likelihood of heterogeneity in outcomes within these groups. The threshold for hospital admission may be lower among individuals with a recent history of COVID-19 than in the general population, and rates of diagnoses in general may have decreased as an indirect result of the pandemic, particularly among people who were not admitted to hospital with COVID-19. We did not have access to testing data and thus were unable to remove individuals infected with COVID-19 who did not require hospitalisation from our control group. Moreover, our results are unlikely to fully capture the lived experience of individuals living with PCS who were possibly asymptomatic and untested at the time of infection. Multi-organ manifestations beyond the acute phase of COVID-19 infection have been identified in non-hospitalised individuals,[23] who were beyond the scope of our study, and we did not capture symptoms such as fatigue, disturbance of taste and smell, and anxiety, which have been widely reported by individuals with suspected PCS.[21]

## Conclusions

Individuals discharged from hospital following acute COVID-19 face elevated rates of mortality, readmission and multi-organ dysfunction compared with the background levels that exist for these individuals, and the relative increase in risk is neither confined to the elderly nor uniform across ethnic groups. Urgent research is required to further understand the risk factors for PCS, so that treatment provision can be better targeted to demographically and clinically at-risk populations.

## Data Availability

In accordance with NHS Digital's Information Governance (IG) requirements, it is not possible for the study data to be shared.

## Footnotes

### Acknowledgements

The authors thank staff at NHS Digital for facilitating access to the data and providing valuable guidance as to its quality and usage, and Neil Bannister and Myer Glickman at the Office for National Statistics (ONS) for their input throughout the study. KK is supported by the National Institute for Health Research (NIHR) Applied Research Collaboration East Midlands (ARC EM) and the NIHR Leicester Biomedical Research Centre (BRC).

### Contributors

DA, KK, VN, BH and AB conceptualised and designed the study. DA and TM prepared the study data and performed statistical analysis. All authors contributed to interpretation of the results. KK, VN, BH, ID and AB contributed to critical revision of the manuscript. All authors approved the final manuscript. The corresponding author attests that all listed authors meet authorship criteria and that no others meeting the criteria have been omitted.

### Competing interests

All authors have completed the ICMJE uniform disclosure form at www.icmje.org/coi_disclosure.pdf and declare: no support from any organisation for the submitted work; no financial relationships with any organisations that might have an interest in the submitted work in the previous three years; KK is Chair of the Ethnicity Subgroup of the Independent Scientific Advisory Group for Emergencies (SAGE), a member of Independent SAGE, a Trustee of the South Asian Health Foundation (SAHF), and Director of the University of Leicester Centre for Black Minority Ethnic Health; and AB is a Trustee of the SAHF, and has received a research grant unrelated to the current work from AstraZeneca.

### Ethics approval

NHS Digital approved access to the data for this study following a favourable recommendation from its Independent Group Advising on the Release of Data (IGARD). Ethical approval was obtained from the National Statistician’s Data Ethics Advisory Committee [NSDEC(20)12].

### Funding source

There was no external funding for this study.

## Supplementary tables

**Supplementary Table 1.** Description of variables used to match COVID-19 and control patients.

| Variable | Categorisation |
| --- | --- |
| Age group | <50 years, 50-69 years, ≥70 years |
| Sex | Female, male |
| Ethnic group | Asian, Black, White, Mixed/Other, Unknown |
| Region of residence | North (North East, North West, Yorkshire and the Humber), South (East of England, London, South East, South West), Midlands (East Midlands, West Midlands), Unknown |
| IMD quintile | 1 (most deprived), 2, 3, 4, 5 (least deprived), Unknown |
| Smoking status | Current, Former, Never/Unknown |
| BMI group | <25 kg/m<sup>2</sup> or Unknown, 25 kg/m<sup>2</sup> to <30 kg/m<sup>2</sup>, ≥30 kg/m<sup>2</sup> |
| History of hypertension | Yes, No |
| History of MACE | Yes, No |
| History of respiratory disease | Yes, No |
| History of CKD stages 3-5 | Yes, No |
| History of CLD | Yes, No |
| History of diabetes | Yes, No |
| History of cancer | Yes, No |

Table notes: BMI: body mass index; CKD: chronic kidney disease; CLD: chronic liver disease; IMD: Index of Multiple Deprivation; MACE: major adverse cardiovascular event (a composite of heart failure, myocardial infarction, stroke, and arrhythmia).

**Supplementary Table 2.**
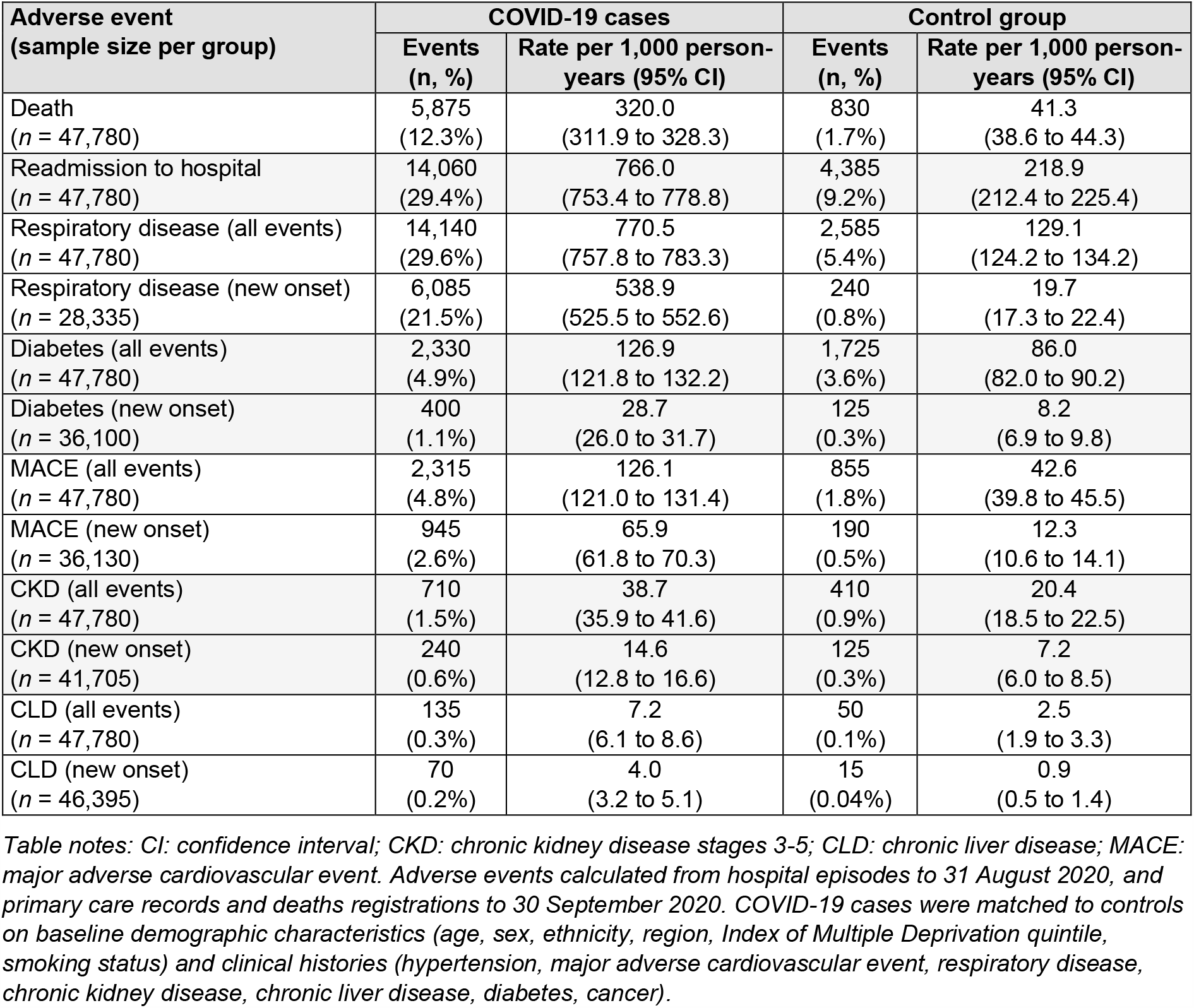
Counts and rates of adverse events contrasting individuals with COVID-19 in England discharged from hospital by 31 August 2020 with matched controls

| Adverse event<br>(sample size per group) | COVID-19 cases |  | Control group |  |
| --- | --- | --- | --- | --- |
|  | Events<br>(n, %) | Rate per 1,000 person-years (95% CI) | Events<br>(n, %) | Rate per 1,000 person-years (95% CI) |
| Death<br>(n = 47,780) | 5,875<br>(12.3%) | 320.0<br>(311.9 to 328.3) | 830<br>(1.7%) | 41.3<br>(38.6 to 44.3) |
| Readmission to hospital<br>(n = 47,780) | 14,060<br>(29.4%) | 766.0<br>(753.4 to 778.8) | 4,385<br>(9.2%) | 218.9<br>(212.4 to 225.4) |
| Respiratory disease (all events)<br>(n = 47,780) | 14,140<br>(29.6%) | 770.5<br>(757.8 to 783.3) | 2,585<br>(5.4%) | 129.1<br>(124.2 to 134.2) |
| Respiratory disease (new onset)<br>(n = 28,335) | 6,085<br>(21.5%) | 538.9<br>(525.5 to 552.6) | 240<br>(0.8%) | 19.7<br>(17.3 to 22.4) |
| Diabetes (all events)<br>(n = 47,780) | 2,330<br>(4.9%) | 126.9<br>(121.8 to 132.2) | 1,725<br>(3.6%) | 86.0<br>(82.0 to 90.2) |
| Diabetes (new onset)<br>(n = 36,100) | 400<br>(1.1%) | 28.7<br>(26.0 to 31.7) | 125<br>(0.3%) | 8.2<br>(6.9 to 9.8) |
| MACE (all events)<br>(n = 47,780) | 2,315<br>(4.8%) | 126.1<br>(121.0 to 131.4) | 855<br>(1.8%) | 42.6<br>(39.8 to 45.5) |
| MACE (new onset)<br>(n = 36,130) | 945<br>(2.6%) | 65.9<br>(61.8 to 70.3) | 190<br>(0.5%) | 12.3<br>(10.6 to 14.1) |
| CKD (all events)<br>(n = 47,780) | 710<br>(1.5%) | 38.7<br>(35.9 to 41.6) | 410<br>(0.9%) | 20.4<br>(18.5 to 22.5) |
| CKD (new onset)<br>(n = 41,705) | 240<br>(0.6%) | 14.6<br>(12.8 to 16.6) | 125<br>(0.3%) | 7.2<br>(6.0 to 8.5) |
| CLD (all events)<br>(n = 47,780) | 135<br>(0.3%) | 7.2<br>(6.1 to 8.6) | 50<br>(0.1%) | 2.5<br>(1.9 to 3.3) |
| CLD (new onset)<br>(n = 46,395) | 70<br>(0.2%) | 4.0<br>(3.2 to 5.1) | 15<br>(0.04%) | 0.9<br>(0.5 to 1.4) |

Table notes: CI: confidence interval; CKD: chronic kidney disease stages 3-5; CLD: chronic liver disease; MACE: major adverse cardiovascular event. Adverse events calculated from hospital episodes to 31 August 2020, and primary care records and deaths registrations to 30 September 2020. COVID-19 cases were matched to controls on baseline demographic characteristics (age, sex, ethnicity, region, Index of Multiple Deprivation quintile, smoking status) and clinical histories (hypertension, major adverse cardiovascular event, respiratory disease, chronic kidney disease, chronic liver disease, diabetes, cancer).

**Supplementary Table 3.** Counts and rates of adverse events contrasting individuals with COVID-19 in England discharged from hospital by 31 August 2020 with matched controls, stratified by ICU versus non-ICU admission

| Group<br>(sample size) | Adverse event | COVID-19 cases |  | Control group |  |
| --- | --- | --- | --- | --- | --- |
|  |  | Events<br>(n, %) | Rate per 1,000 person-years (95% CI) | Events<br>(n, %) | Rate per 1,000 person-years (95% CI) |
| ICU patients<br>(n = 4,745) | Death | 420<br>(8.8%) | 237.9<br>(215.7 to 261.8) | 20<br>(0.5%) | 11.4<br>(7.2 to 17.3) |
|  | Readmission | 1,235<br>(26.0%) | 701.2<br>(662.6 to 741.4) | 260<br>(5.5%) | 134.8<br>(118.9 to 152.2) |
|  | Respiratory<br>disease | 1,620<br>(34.1%) | 918.6<br>(874.4 to 964.5) | 100<br>(2.1%) | 53.1<br>(43.3 to 64.4) |
|  | Diabetes | 335<br>(7.1%) | 190.2<br>(170.4 to 211.7) | 175<br>(3.6%) | 90.0<br>(77.1 to 104.5) |
|  | MACE | 175<br>(3.6%) | 98.2<br>(84.1 to 114.0) | 30<br>(0.6%) | 15.1<br>(10.1 to 21.7) |
|  | CKD | 65<br>(1.3%) | 35.8<br>(27.5 to 45.8) | 20<br>(0.4%) | 9.4<br>(5.6 to 14.8) |
| Non-ICU<br>patients<br>(n = 43,035) | Death | 5,455<br>(12.7%) | 328.8<br>(320.1 to 337.6) | 805<br>(1.9%) | 44.5<br>(41.5 to 47.7) |
|  | Readmission | 12,825<br>(29.8%) | 772.9<br>(759.5 to 786.4) | 4,125<br>(9.6%) | 227.8<br>(220.9 to 234.8) |
|  | Respiratory<br>disease | 12,525<br>(29.1%) | 754.7<br>(741.6 to 768.1) | 2,485<br>(5.8%) | 137.2<br>(131.8 to 142.7) |
|  | Diabetes | 1,995<br>(4.6%) | 120.2<br>(115.0 to 125.6) | 1,550<br>(3.6%) | 85.6<br>(81.4 to 90.0) |
|  | MACE | 2,140<br>(5.0%) | 129.1<br>(123.7 to 134.7) | 825<br>(1.9%) | 45.5<br>(42.4 to 48.7) |
|  | CKD | 645<br>(1.5%) | 39.0<br>(36.0 to 42.1) | 390<br>(0.9%) | 21.6<br>(19.5 to 23.8) |

Table notes: CI: confidence interval; CKD: chronic kidney disease stages 3-5; ICU: intensive care unit; MACE: major adverse cardiovascular event. Adverse events calculated from hospital episodes to 31 August 2020, and primary care records and deaths registrations to 30 September 2020. COVID-19 cases were matched to controls on baseline demographic characteristics (age, sex, ethnicity, region, Index of Multiple Deprivation quintile, smoking status) and clinical histories (hypertension, major adverse cardiovascular event, respiratory disease, chronic kidney disease, chronic liver disease, diabetes, cancer).

**Supplementary Table 4.** Rates and rate ratios of adverse events contrasting individuals with COVID-19 in England discharged from hospital by 31 August 2020 with matched controls, stratified by demographic factors

| Group (sample size) | Adverse event | Rate per 1,000 person-years (95% CI) |  | Rate ratio (95% CI) |
| --- | --- | --- | --- | --- |
|  |  | COVID-19 cases | Control group |  |
| Sex: Male (n = 26,245) | Death | 318.4 (307.5 to 329.6) | 38.5 (34.9 to 42.3) | 8.3 (7.5 to 9.2) |
|  | Readmission | 757.5 (740.7 to 774.7) | 214.3 (205.8 to 223.1) | 7.2 (6.5 to 8.0) |
|  | Respiratory | 780.9 (763.8 to 798.3) | 127.3 (120.7 to 134.1) | 14.1 (11.0 to 18.3) |
|  | Diabetes | 142.4 (135.2 to 150.0) | 95.6 (90.0 to 101.6) | 7.7 (7.1 to 8.3) |
|  | MACE | 130.7 (123.7 to 137.9) | 43.0 (39.2 to 47.1) | 7.6 (7.0 to 8.2) |
|  | CKD | 43.7 (39.7 to 48.0) | 20.5 (17.9 to 23.4) | 10.0 (7.3 to 13.8) |
| Sex: Female (n = 21,535) | Death | 322.0 (309.9 to 334.5) | 44.8 (40.6 to 49.4) | 3.5 (3.4 to 3.7) |
|  | Readmission | 776.4 (757.5 to 795.7) | 224.5 (214.8 to 234.5) | 3.5 (3.3 to 3.6) |
|  | Respiratory | 757.6 (738.9 to 776.6) | 131.4 (124.0 to 139.1) | 4.4 (4.1 to 4.6) |
|  | Diabetes | 107.9 (101.0 to 115.3) | 74.2 (68.6 to 80.0) | 3.2 (3.1 to 3.3) |
|  | MACE | 120.6 (113.2 to 128.3) | 42.0 (37.9 to 46.5) | 3.3 (3.2 to 3.5) |
|  | CKD | 32.5 (28.8 to 36.7) | 20.3 (17.5 to 23.5) | 4.4 (4.0 to 4.9) |
| Age group: <70 years (n = 25,955) | Death | 86.2 (80.7 to 91.9) | 6.1 (4.7 to 7.7) | 6.1 (5.8 to 6.5) |
|  | Readmission | 556.6 (542.6 to 570.8) | 127.0 (120.5 to 133.8) | 5.8 (5.4 to 6.1) |
|  | Respiratory | 615.8 (601.1 to 630.8) | 58.5 (54.1 to 63.1) | 10.5 (9.7 to 11.4) |
|  | Diabetes | 123.9 (117.4 to 130.7) | 73.2 (68.2 to 78.4) | 4.6 (4.3 to 4.8) |
|  | MACE | 55.7 (51.4 to 60.4) | 13.0 (11.0 to 15.3) | 5.2 (5.0 to 5.5) |
|  | CKD | 24.4 (21.6 to 27.5) | 6.9 (5.4 to 8.6) | 11.4 (9.8 to 13.3) |
| Age group: ≥70 years (n = 21,825) | Death | 658.4 (640.1 to 677.0) | 85.6 (79.6 to 91.9) | 1.5 (1.4 to 1.6) |
|  | Readmission | 1,069.0 (1,045.7 to 1,092.6) | 334.2 (322.3 to 346.5) | 1.5 (1.3 to 1.6) |
|  | Respiratory | 994.2 (971.7 to 1,017.0) | 217.8 (208.2 to 227.7) | 1.7 (1.6 to 1.8) |
|  | Diabetes | 131.3 (123.2 to 139.8) | 102.1 (95.6 to 109.0) | 1.3 (1.2 to 1.4) |
|  | MACE | 227.9 (217.3 to 239.0) | 79.7 (74.0 to 85.8) | 1.4 (1.3 to 1.5) |
|  | CKD | 59.3 (53.9 to 65.1) | 37.4 (33.5 to 41.6) | 1.5 (1.3 to 1.7) |
| Ethnic group: White (n = 34,355) | Death | 403.4 (392.5 to 414.5) | 53.1 (49.4 to 57.0) | 3.0 (2.7 to 3.4) |
|  | Readmission | 876.8 (860.7 to 893.2) | 263.6 (255.3 to 272.2) | 2.9 (2.5 to 3.2) |
|  | Respiratory | 861.1 (845.1 to 877.3) | 164.5 (157.9 to 171.3) | 4.3 (3.6 to 5.2) |
|  | Diabetes | 119.6 (113.6 to 125.7) | 85.5 (80.8 to 90.4) | 2.9 (2.6 to 3.1) |
|  | MACE | 153.4 (146.7 to 160.3) | 53.9 (50.2 to 57.9) | 2.8 (2.6 to 3.1) |
| Ethnic group: Non-White (n = 8,315) | Death | 126.6 (115.0 to 139.0) | 12.7 (9.3 to 16.9) | 2.1 (1.8 to 2.5) |
|  | Readmission | 553.7 (529.2 to 579.0) | 126.1 (114.8 to 138.2) | 1.6 (1.3 to 1.9) |
|  | Respiratory | 567.8 (543.0 to 593.4) | 49.9 (42.9 to 57.8) | 3.5 (2.7 to 4.6) |
|  | Diabetes | 184.1 (170.1 to 198.9) | 121.1 (110.1 to 133.0) | 1.6 (1.4 to 1.8) |
|  | MACE | 68.2 (59.8 to 77.4) | 18.8 (14.6 to 23.8) | 1.7 (1.5 to 2.0) |

Table notes: CI: confidence interval; CKD: chronic kidney disease stages 3-5; MACE: major adverse cardiovascular event. Adverse events calculated from hospital episodes to 31 August 2020, and primary care records and deaths registrations to 30 September 2020. COVID-19 cases were matched to controls on baseline demographic characteristics (age, sex, ethnicity, region, Index of Multiple Deprivation quintile, smoking status) and clinical histories (hypertension, major adverse cardiovascular event, respiratory disease, chronic kidney disease, chronic liver disease, diabetes, cancer). ). Rates and rate ratios for CKD could not be stratified by ethnic group due to insufficient event counts in the control group.

## Notes

### Author Declarations

Ethical approval was obtained from the National Statistician's Data Ethics Advisory Committee [NSDEC(20)12].

## References

[1] Ward H, Atchison C, Whitaker M, et al. Antibody prevalence for SARS-CoV-2 in England following first peak of the pandemic: REACT2 study in 100,000 adults. 2020. https://www.medrxiv.org/content/10.1101/2020.08.12.20173690v2

[2] Banerjee A, Pasea L, Harris S, et al. Estimating excess 1-year mortality associated with the COVID-19 pandemic according to underlying conditions and age: a population-based cohort study. Lancet 2020;395:1715–25. doi: https://doi.org/10.1016/S0140-6736(20)30854-0

[3] Clift A, Coupland C, Keogh R, et al. Living risk prediction algorithm (QCOVID) for risk of hospital admission and mortality from coronavirus 19 in adults: national derivation and validation cohort study. BMJ 2020;371:m3731. doi: https://doi.org/10.1136/bmj.m3731

[4] Williamson E, Walker A, Bhaskaran K, et al. Factors associated with COVID-19-related death using OpenSAFELY. Nature 2020;548:430–36. doi: https://doi.org/10.1038/s41586-020-2521-4

[5] World Health Organisation. Clinical management of severe acute respiratory infection (SARI) when COVID-19 disease is suspected: interim guidance, 13 March 2020. 2020. https://apps.who.int/iris/handle/10665/331446

[6] Puntmann V, Ludovica Carer M, Wieters I, et al. Outcomes of Cardiovascular Magnetic Resonance Imaging in Patients Recently Recovered From Coronavirus Disease 2019 (COVID-19). JAMA Cardiol 2020;5:1265–73. doi: 10.1001/jamacardio.2020.3557

[7] Tabary M, Khanmohammadi S, Araghi F, et al. Pathologic features of COVID-19: A concise review. Pathol Res Pract 2020;216:153097. doi: 10.1016/j.prp.2020.153097

[8] Alqahtani S, Schattenberg J. Liver injury in COVID-19: The current evidence. United European Gastroenterol J 2020;8:509–19. doi: 10.1177/2050640620924157

[9] Somasundaram N, Ranathunga I, Ratnasamy V, et al. The Impact of SARS-Cov-2 Virus Infection on the Endocrine System. J Endocr Soc 2020;4:bvaa082. doi: 10.1210/jendso/bvaa082

[10] Farouk S, Fiaccadori E, Cravedi P, et al. COVID-19 and the kidney: what we think we know so far and what we don’t. J Nephrol 2020;33:1213–18. doi: https://doi.org/10.1007/s40620-020-00789-y

[11] Ball S, Banerjee A, Berry C, et al. Monitoring indirect impact of COVID-19 pandemic on services for cardiovascular diseases in the UK. Heart 2020;106:1890–97. doi: 10.1136/heartjnl-2020-317870

[12] Lai A, Pasea L, Banerjee A, et al. Estimated impact of the COVID-19 pandemic on cancer services and excess 1-year mortality in people with cancer and multimorbidity: near real-time data on cancer care, cancer deaths and a population-based cohort study. BMJ Open 2020;10:e043828. doi: 10.1136/bmjopen-2020-043828

[13] Katsoulis M, Gomes M, Lai A, et al. Estimating the effect of reduced attendance at emergency departments for suspected cardiac conditions on cardiac mortality during the COVID-19 pandemic. 2020. https://www.ahajournals.org/doi/10.1161/CIRCOUTCOMES.120.007085

[14] National Institute for Health and Care Excellence. COVID-19 rapid guideline: managing the long-term effects of COVID-19. 2020. https://www.nice.org.uk/guidance/ng188

[15] Greenhalgh T, Knight M, A’Court C, et al. Management of post-acute covid-19 in primary care. BMJ 2020;370:m3026. doi: 10.1136/bmj.m3026

[16] National Institute for Health Research. Themed Review: Living with Covid19. 2020. https://evidence.nihr.ac.uk/themedreview/living-with-covid19

[17] Austin, P. Balance diagnostics for comparing the distribution of baseline covariates between treatment groups in propensity-score matched samples. Stat Med 2009;28:3083–107. doi: 10.1002/sim.3697

[18] Fraser, E. Long term respiratory complications of covid-19. BMJ 2020;370:m3001. doi: https://doi.org/10.1136/bmj.m3001

[19] Becker, R. Anticipating the long-term cardiovascular effects of COVID-19. J Thromb Thrombolysis 2020;50:512–24. doi: 10.1007/s11239-020-02266-6

[20] Rubino F, Amiel S, Zimmet P, et al. New-Onset Diabetes in Covid-19. N Engl J Med 2020;383:789–90. doi: 10.1056/NEJMc2018688

[21] Michelen M, Manoharan L, Elkheir N, et al. Characterising long-term covid-19: a rapid living systematic review. 2020. https://www.medrxiv.org/content/10.1101/2020.12.08.20246025v1

[22] Donnelly J, Qing Wang X, Iwashyna T. Readmission and Death After Initial Hospital Discharge Among Patients With COVID-19 in a Large Multihospital System. JAMA 2020;e2021465. doi: 10.1001/jama.2020.21465

[23] Dennis A, Wamil M, Kapur S, et al. Multi-organ impairment in low-risk individuals with long COVID. 2020. https://www.medrxiv.org/content/10.1101/2020.10.14.20212555v1

[24] McCarthy C, Murphy S, Jones-O’Connor M, et al. Early clinical and sociodemographic experience with patients hospitalized with COVID-19 at a large American healthcare system. Lancet 2020;26:100504. doi: 10.1016/j.eclinm.2020.100504

[25] Xie Y, Bowe B, Maddukuri G. Comparative evaluation of clinical manifestations and risk of death in patients admitted to hospital with covid-19 and seasonal influenza: cohort study. BMJ 2020;371:m4677. doi: 10.1136/bmj.m4677

[26] Liu D, Zhang W, Pan F, et al. The pulmonary sequalae in discharged patients with COVID-19: a short-term observational study. Respir Res 2020;21:125. doi: 10.1186/s12931-020-01385-1

[27] Rajpal S, Tong M, Borchers J, et al. Cardiovascular Magnetic Resonance Findings in Competitive Athletes Recovering From COVID-19 Infection. JAMA Cardiol 2020;e204916. doi: 10.1001/jamacardio.2020.4916

[28] UK Government. Coronavirus (COVID-19) in the UK: daily update. https://coronavirus.data.gov.uk

[29] Campbell H, Hotchkiss R, Bradshaw N, et al. Integrated care pathways. BMJ 1998;316:133–37. doi: 10.1136/bmj.316.7125.133

